# Enhancing Histopathological Colorectal Cancer Image Classification by using Convolutional Neural Network

**DOI:** 10.1101/2021.03.17.21253390

**Authors:** Radwan Al.Shawesh, Yi Xiang Chen

## Abstract

Colorectal cancer (CRC) also known as bowl cancer is one of the leading death causes worldwide. Early diagnosis has become vital for a successful treatment. Now days with the new advancements in Convolutional Neural networks (CNNs) it’s possible to classify different images of CRC into different classes. Today It is crucial for physician to take advantage of the new advancement’s in deep learning, since classification methods are becoming more and more accurate and efficient. In this study, we introduce a method to improve the classification accuracy from previous studies that used the National Center for Tumor diseases (NCT) data sets. We adapt the ResNet-50 model in our experiment to classify the CRC histopathological images. Furthermore, we utilize transfer learning and fine-tunning techniques to improve the accuracy. Our Experiment results show that ResNet_50 network is the best CNN architecture so far for classifying CRC histopathological images on the NCT Biobank open source dataset. In addition to that using transfer learning allow us to obtain 97.7% accuracy on the validation dataset, which is better than all previous results we found in literature.

## Introduction

Cancer is one of the leading death causes worldwide. According to the World Health Organization (WHO) GLOBOCAN database, there are 1,849,518 estimated new Colorectal Cancer (CRC) cases and 880,792 CRC-related deaths in 2018 [1]. Thus making CRC the fourth most commonly diagnosed cancer and the third leading cause of cancer death in the world. It was also found CRC is 3-4 times more common in developed than in developing nations [2]. For instance, the American cancer society, found in a study that CRC is the second leading cause of cancer-related deaths in men and women combined [3]. As a result diagnosis of CRC has become extremely important especially in early stages. Today biopsy is the most reliable way to make sure whether an area of the body has cancer [4]. The process involves the removal of a small amount of tissue for examination under a microscope, and then staining them. After that, histopthological slides are digitized resulting in images that pathologist can analyze [5].

However, this technique might get replaced by new modern techniques since the traditional way of diagnosing cancer via a biopsy test is time-consuming, and requires the prior knowledge of professionals. In addition, the result of the diagnosis may vary according to the level of experience of the pathologist [4]. Now with the new advancement in neural networks and image classifications, Convolutional neural networks (CNNs) have been used to analyze histopathological images and classify cancer tissues [6]. CNNs have been one of the hottest topics over the last few years on classifying images and making decisions. CNN is a common type of machine learning that is based on how our nervous system learns and operates. These Neural networks uses self-adaptive algorithms that learn from data to implement what they learn and make decisions [7].

Since 2012, when the groundbreaking AlexNet was first introduced by Alex Krizevsky [8], CNN has contributed in many breakthroughs in computer vision, and has revolutionized the image analysis field. Complex visual tasks can be efficiently accomplished by CNN that can learn to differentiate between objects based on features learned from training data. The application of CNN range widely, from facial recognition [9], document analysis [10], traffic sign analysis [11] to understanding and forecasting the climate trends [12]. While in the context of medical imaging, medical images in general [13–16], and histopathological images in specific [17 –23], became important application of deep learning. It is now possible to use CNN to categorize histopathological biopsy images based on the detection of features and patterns within the image itself. Moreover, CNN have the ability to automatically extract features from data without any prior knowledge. This can solve the problem of traditional feature extraction techniques which are low in efficiency and often produce poor results [24].

While there are many notable achievements in the area of classifying histology images, there wasn’t many researches done on classifying the tissues of CRC [25, 26]. Moreover, many results from several studies still suffer from having unreliable accuracy results. This is mainly due to the complex structures and variability within a class of histology images.

In this study, our goal is to fill these gaps in the context of CRC. We use an advanced deep convolutional neural network, like the ResNet_50 [27], combined with transfer learning and finetuning techniques [28, 29] to classify the histopathological images of CRC. We use two large data sets of histology images from two different sources in order to evaluate the results of our CNN. We also be able to compare our results with other studies since the datasets we use were open source and has been used before to conduct similar experiments to ours.

## Materials and methods

### Dataset

The two datasets used in this article was published by Kather, J. N. in 2019 [30]. The first dataset National Center for Tumor Diseases (NCT-CRC-HE-100K) consist of 100K of non-overlapping images manually extracted from N=86 HE stained human cancer tissue slides from the NCT biobank (National Center for Tumor diseases) and the UMM pathology archive (University Medical Center Mannheim). While the second dataset Colorectal Cacner-Validation-Histology-7K (CRC-VAL-HE-7K) consist of 7180 images extracted from 50 patients with colorectal adenocarcinoma and were used to create a dataset that does not overlap with patients in the first dataset. All Images in both dataset are 224×224 pixels at 0.5 MPP (Micron per pixel). Furthermore, in order to eliminate the inconsistencies in the staining process and to improve the classification result, all images are color-normalized by the author with the Macenko method [31].

We use the NCT-CRC-HE-100K as our training set and the CRC-VAL-HE-7K as our validation set. Each set consist of 9 classes of tissue: Adipose (ADI), background (BACK), debris (DEB), lymphocytes (LYM), mucus (MUC), smooth muscle (MUS), normal colon mucosa (NORM), cancer-associated stroma (STR), colorectal adenocarcinoma epithelium (TUM) [30]. Table 1 shows a description of the NCT biobank database. The training and validation dataset can be accessed through the link http://dx.doi.org/10.5281/zenodo.1214456.

**Table 1.**
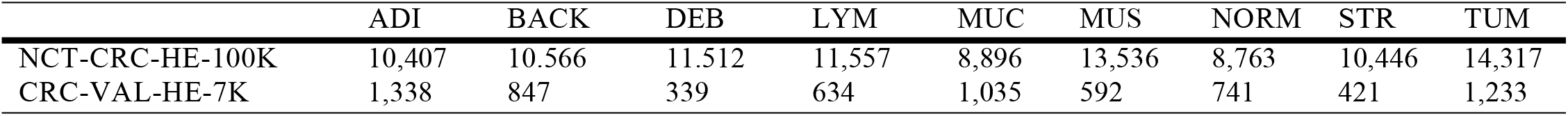
Image distribution of different tissue classes in the NCT database

Since the input size requirement of ResNet_50 network used in this study matches the size of the images in the database it won’t be necessary to transform the images to a new input size. In addition to that no preprocessing methods been used to transform the images in any way.

### Classification Analysis

This section will introduce the method used to classify the histology images of CRC using the deep learning model of ResNet_50 [27].

#### CNN structure for classification

The ResNet_50 [27] proposed by He et al. in 2015 is adopted in our experiment. ResNet_50 is equipped with residual connections and is 50 layers deep, meaning its much deeper and superior to many other networks. This is mainly due to the residual units which allows the neural network to train more layers whileavoiding deterioration of the network gradient. ResNet first introduced the concept of skip connection. It can be seen from Fig 1 that the network consist of 5 stages each with a convolution and Identity block as shown in Fig 2. Those blocks act as highways were the network is able to skip layers and take shortcuts in order to improve performance.

**Figure 1.**
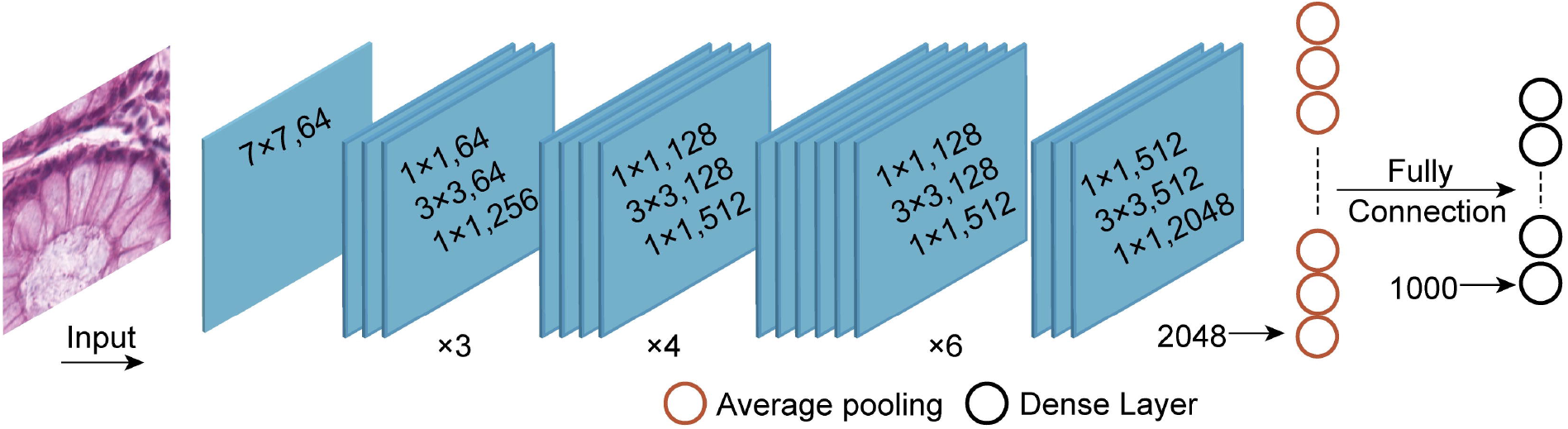
The network structure ResNet_50 [27]

**Figure 2.**
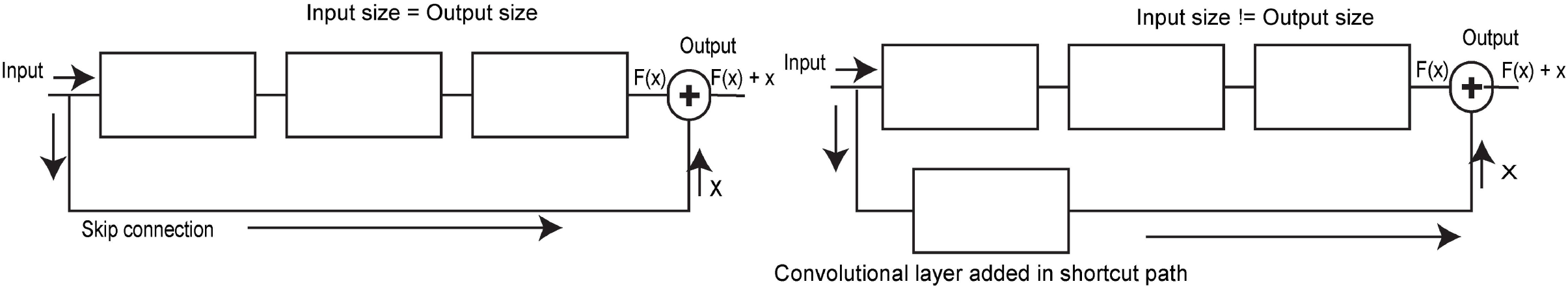
Residual learning. The image in the left shows the identity block, and the image in the right shows the convolutional block [27]

#### Training ResNet_50 via Transfer learning

Transfer learning [28] is used in this study to train the model of ResNet_50 [27]. We decided to use transfer learning for mainly two reasons: first reasons is to avoid training from scratch since training a neural network from scratch and achieving great results is almost impossible for most people, this is due to the fact that many state of the art neural networks the likes of ResNet_50 and many others were trained on ImageNet dataset. This dataset contains millions of training, testing, and validation images. Moreover, it has a total of 1,000 different categories. Since our dataset is incomparable to ImageNet in terms of its size, we decided to adopt a model that was already trained on the ImageNet database and use the knowledge it has acquired as pre-training for our specific research task. The second reason is training a model the likes of ResNet_50 from scratch is time-consuming and requires huge computational power this might not be worth it if your target goal is just to achieve excellent results in your specific task.

We can see from Fig 3 which shows our modified network structure of ResNet_50, the model consist of frozen and trainable layers. The frozen layers are colored in blue as shown in the figure. While the last few layers and the fully connected layers are set to be trainable. The input is an 224×224 image, followed by a single convolutional layer that reshape the input image into half of its original size. After that comes 4 stacks or blocks of layers each containing multiple layers. In the end of the network we have the average pooling layer followed by the fully connected layer which uses SoftMax function to output the results and classify the images into 9 different classes.

**Figure 3.**
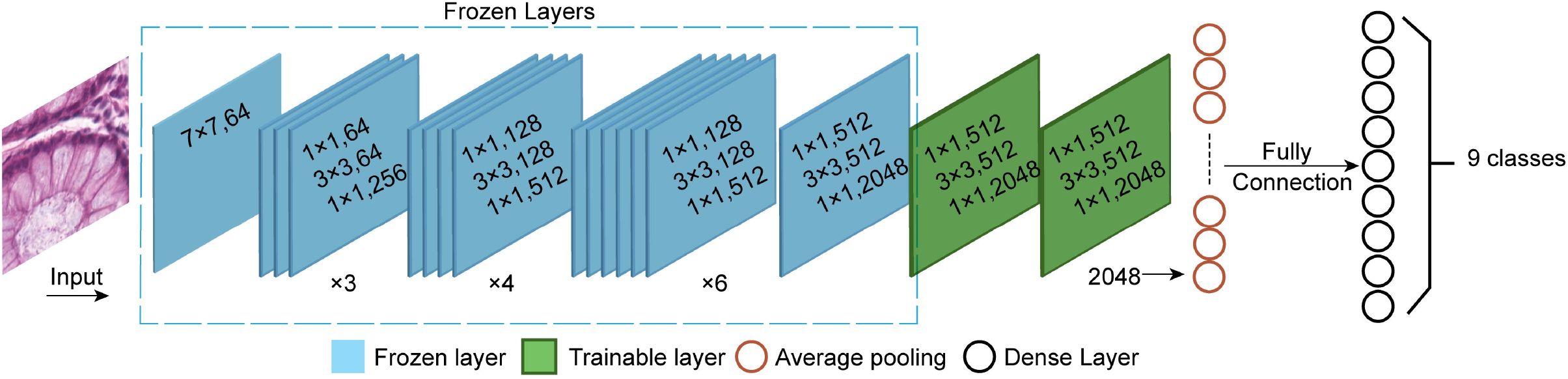
The ResNet_50 fine-tunned network structure for transfer learning

### Evaluationcriteria

In order to be able to compare our results with results from other studies, the performance of the network was evaluated in terms of accuracy. We used the entire CRC-VAL-HE-7K validation set to evaluate our result, and measured the average accuracy across the 9 tissue classes.

## Results

In this section we show our classification results on the 100K histopathological images of CRC from the NCT biobank database, and provide some analyses and discussions of the results. In addition to this, we present a comparison between our experiment results and the results made by other studies.

### Experiment

We train the network on a desktop workstation using a single Nvidia RTX2070SUPER GPU. The training process is conducted as follows: We first download the model and parameters of ResNet_50 network trained on the ImageNet dataset. Then, we froze all the parameters before the last few layers of the network. After that we fine-tune the fully connected layer and change the numbers of neurons of the dense layer as 9 for multi-class classification. Finally, we train the unfrozen layers of the network until we archive the best results.

We use TensorFlow deep learning framework to develop and implement the entire classification process. Since we only have 8Gb of memory to work with we set the batch size to 10. We chose the Adam (adaptive moment estimation) algorithm as our optimizer cause it is fast and reliable. As for the learning rate we use the default Adam learning rate since it can handle learning rate optimizations and for the output we decided to use SoftMax as our output function. When it come to data augmentation, we decided to train on the raw data in order to save time.

### Classification Results

We use the NCT biobank dataset to train our fine-tunned ResNet_50 via transfer learning and test the classification performance in an independent set of 7,180 images from a variety of patients. We did a multi-class classification diagnosis study on the histopathological images of colorectal cancer by using ResNet_50 with transfer learning and fine-tunning techniques. The test results of our multi-class classification of histopathological images of colorectal cancer are shown in Fig 4.

The training accuracy was close to 99%, while the validation accuracy reached 97.7%. A high accuracy was obtained in all of the 9 classes of CRC tissues. While the results in Fig 5 shows that the value of the loss function decreases fast and smoothly converges to small value during the training test.

**Figure 4.**
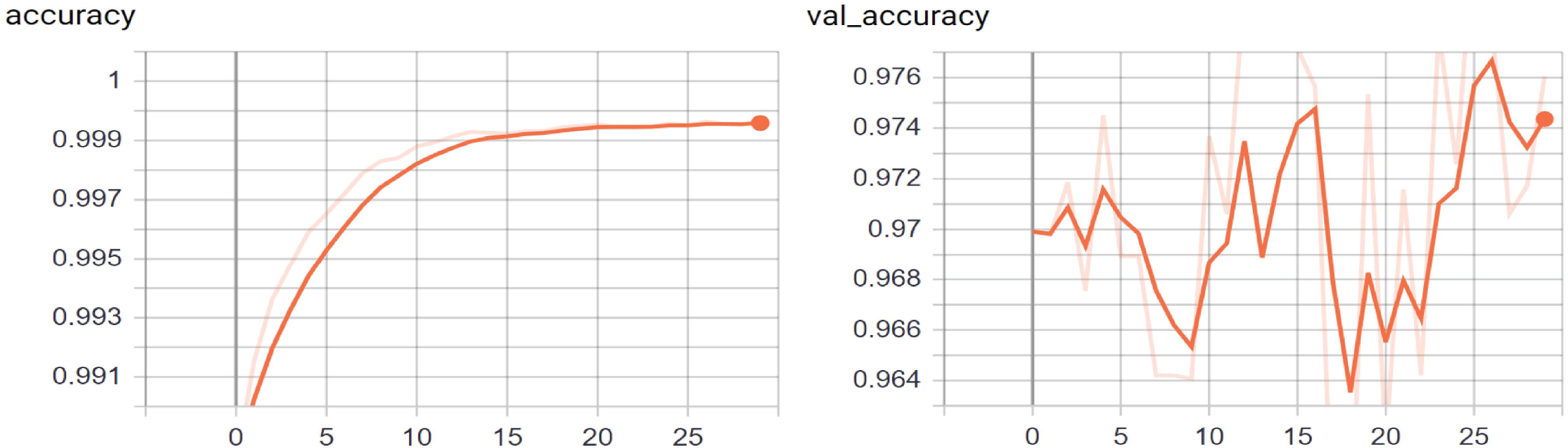
Training accuracy and validation accuracy of the CRC histology images classification results

**Figure 5.**
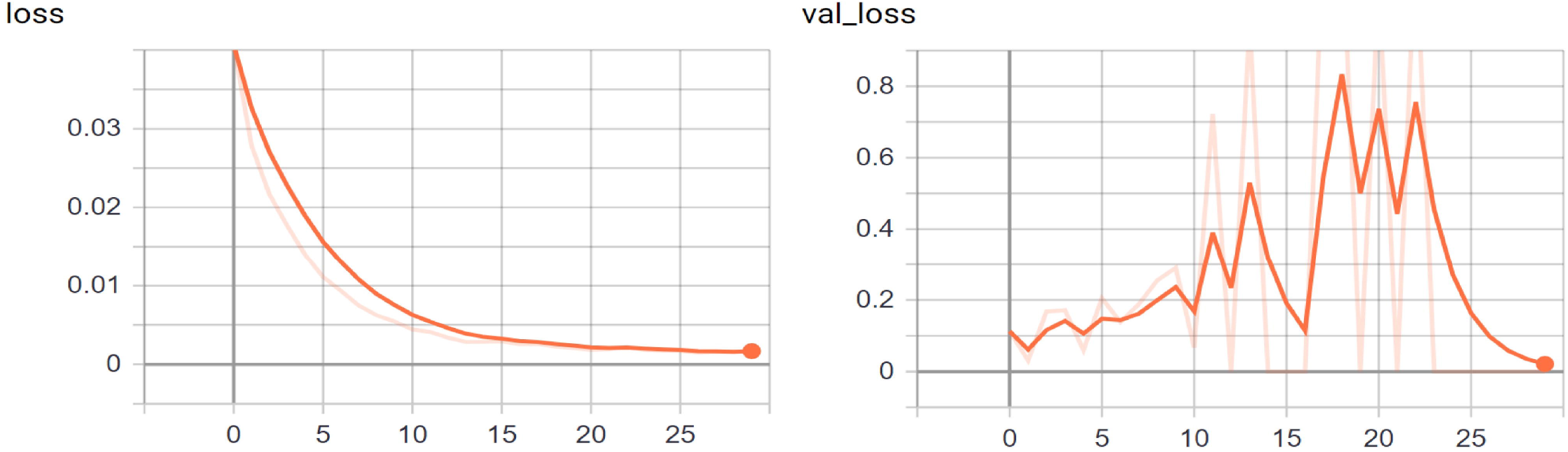
The loss function value during the training and validation of ResNet_50 on the histopathological images of CRC

### Results comparisons

In this subsection we will compare the test results of classifying histopathological images of colorectal cancer using ResNet_50 with transfer learning and fine-tunning techniques. The results will be compared in terms of accuracy since most available studies only used this evaluation criteria. Table 2 shows a comparison between our results and the results made by other studies on the NCT-CRC-HE-100K and CRC-VAL-HE-7K databases. We compared our classification accuracy to the results in [25, 26]. The results tell us that our fine-tunned network have the best accuracy among all of the available studies we found in literature concerning the classification of histopathological images of colorectal cancer on the NCT biobank database. Therefore, the fine-tunned neural network of ResNet_50 with transfer learning is a great option for classifying the histopathological images of CRC.

**Table 2.** CRC multi-class classification comparison between our results and results from other studies that used the NCT-CRC-HE-100K and CRC-VAL-HE-7K databases in their classification. The results of Multitask Resnet18, Resnet35, and Resnet50 are quoted from [26]. The results of VGG19 is quoted from [25].

| Testing accuracy | Fine-tuned<br>Resnet_50 + transfer<br>learning(Ours) | Multitask<br>Resnet18 [26] | Resnet18 [26] | Resnet50 [26] | VGG19 [25] |
| --- | --- | --- | --- | --- | --- |
| NCT-CRC-HE-100K | **99.9** | 98.6 | 98.5 | 98.8 | 98.8 |
| CRC-VAL-HE-7K | **97.7** | 95.0 | 94.4 | 94.2 | 94.3 |

## Conclusion

In this study, we successfully enhanced the classification accuracy of histopathological images of CRC. We adapted the ResNet-50 model in our experiment to classify the CRC histopathological images. Furthermore, we utilized transfer learning and fine-tunning techniques to improve the accuracy. Our adopted network was pretrained on the ImageNet dataset. Then its parameters are frozen. The number of neurons in the dense layer is adjusted accordingly, and the parameters of the dense layer and the last few layers of the main network is re-trained. In this way, the model can be used to classify histopathological images of CRC. Experiment results and the comparison to the results from other studies have shown that ResNet_50 network has the advantage when it comes to extracting features from histopathological images of CRC. In addition to that using transfer learning method allowed us to obtain better results than any previous study found in literature.

## Supporting information

codes and data links

## Data Availability

Yes - all data are fully available without restriction

http://dx.doi.org/10.5281/zenodo.1214456

https://drive.google.com/drive/folders/1Xcu1LPBV5FKoe2-HW6OmgCpw7Uz2WLX5?usp=sharing

## Acknowledgments

The author would like to thank the editors and anonymous reviewers for their constructive comments. The author would also like to thank Kather, Jakob Nikolas, Halama, Niels, & Marx, Alexander who provided the database for NCT-CRC-HE-100K and CRC-VAL-HE-7K. This work was supported in part by the National Key Research and Development Project of China (2018YFB2101300).

