## Supplementary material for "Enhancing Histopathological Colorectal Cancer Image Classification by using Convolutional Neural Network": codes and data links

Project coding can be downloaded from this file: 
https://drive.google.com/drive/folders/1Xcu1LPBV5FKoe2-HW6OmgCpw7Uz2WLX5?usp=sharing

Guide: 
Both the "NCT-CRC-HE-100K" training dataset and "CRC-VAL-HE-7K" should be inside the main folder of the project file
Dataset can be downloaded from the folwing link: http://dx.doi.org/10.5281/zenodo.1214456

The Demo File will validate the CRC-VAL-HE-7K results at the end the overall accuracy and a browser link will appear to classify single images of CRC that satasify the input requirments of the model.
Model file is the training folder
